# A systematic review of aperiodic neural activity in clinical investigations

**DOI:** 10.1101/2024.10.14.24314925

**Authors:** Thomas Donoghue

**Author notes:** Corresponding Author:* Thomas Donoghue. Conflicts of Interest:* The author declares no conflicts of interest. **Project Repository:** The project repository contains code & data related to this project: https://github.com/TomDonoghue/AperiodicClinical. **Abbreviations:** ADHD: attention-deficit hyperactivity disorder; DBS: deep brain stimulation; DOC: disorders of consciousness; DOI: digital object identifier; EEG: electroencephalography; E/I: excitation/inhibition; iEEG: intracranial EEG; MEG: magnetoencephalography; PRISMA: Preferred Reporting Items for Systematic reviews and Meta-Analyses; RNS: random noise stimulation.

## Abstract

Aperiodic neural activity – activity with no characteristic frequency – has increasingly become a common feature of study, including in clinical work. Reports investigating aperiodic activity from patients from a broad range of clinical disorders have sought to evaluate aperiodic activity as a putative biomarker relating to diagnosis or treatment response, and/or as a potential marker of underlying physiological activity. However, there is thus far no clear consensus on if and how aperiodic neural activity relates to clinical disorders. This systematic literature review, following PRISMA guidelines, examines reports of aperiodic activity in electrophysiological recordings from human patients with psychiatric and/or neurological disorders, finding 177 reports across 38 disorders. Results are summarized to evaluate current findings and examine what can be learned as pertains to the analysis, interpretations, and overall utility of aperiodic neural activity in clinical investigations. Aperiodic activity is commonly reported to relate to clinical diagnoses, with 32 of 38 disorders reporting a significant effect in diagnostic and/or treatment related studies. However, there is variation in the consistency of results across disorders, with the heterogeneity of patient groups, disease etiologies, and treatment status arising as common themes. Overall, the current variability of results, potentially confounding covariates, and limitations in current understanding of aperiodic activity suggests further work is needed before aperiodic activity can be established as a potential biomarker and/or marker of underlying pathological physiology. A series of recommendations are proposed, to assist with guiding productive future work on the clinical utility of studying aperiodic neural activity.

## 1. Introduction

There is a long history of using neuro-electrophysiological recordings from methods such as electroencephalography (EEG), magnetoencephalography (MEG), and in some cases, invasive recordings such as intracranial EEG (iEEG) to investigate clinical disorders across psychiatry and neurology (Babiloni et al., 2020; Başar & Güntekin, 2008; Donoghue & Voytek, 2022; Newson & Thiagarajan, 2019). Recently, work exploring different methods for analyzing such data has led to a rapid increase in the popularity of the study of aperiodic neural activity as a feature of interest. Aperiodic activity is defined by a lack of a characteristic frequency, as compared to oscillatory (rhythmic) activity that has a reoccurring pattern. Aperiodic activity can be examined by measuring the aperiodic exponent (equivalently, the spectral slope) from the neural power spectrum (Figure 1A-B). Aperiodic neural activity is a dynamic physiological signal, and has been shown to vary systematically through development and in aging (Stanyard et al., 2024; Voytek et al., 2015), across sleep and wake stages (Ameen et al., 2024; Lendner et al., 2020), and during cognitive tasks (Gyurkovics et al., 2022; Waschke et al., 2021).

**Figure 1.**
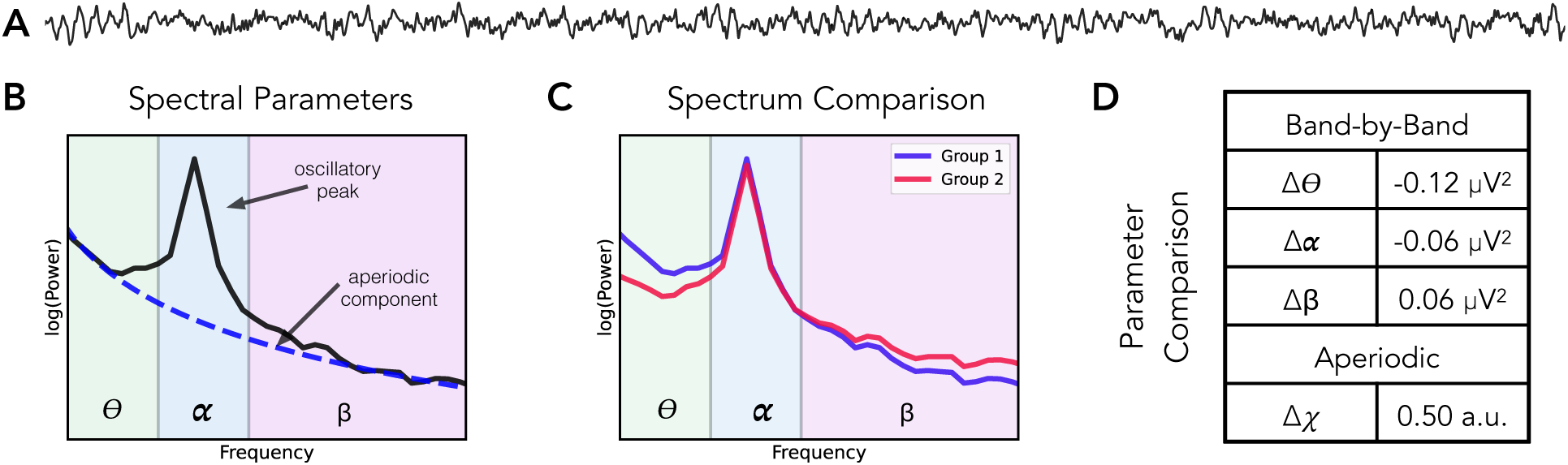
Schematic introducing features of neuro-electrophysiological recordings. **A)** An example (simulated) time series, with a combination of aperiodic activity and a bursty 10 Hz oscillation. **B)** The annotated power spectrum for the signal in (A), showing the estimated power of the signal (black) as well as the measured aperiodic component (blue). Frequency ranges are shaded by typical oscillation band ranges - theta: 3-8 Hz, alpha: 8-13 Hz, and beta: 13-35 Hz. **C)** An example comparison of two power spectra. In this comparison, the difference in the two power spectra was simulated as a change in the aperiodic exponent. **D)** The quantified parameter differences for the example spectra in (C). When measuring power across pre-defined oscillations bands, there is what appears to be a pattern of changes across bands. However, this can be explained by a change in the aperiodic exponent, which is the parameter that was actually changed in this simulation.

Methodologically, a key motivation for measuring aperiodic activity is due to its potential for confounding more traditional measures of oscillatory activity (Figure 1C-D). Specifically, analyses designed to examine oscillatory activity may actually reflect aperiodic activity, which can lead to erroneous interpretations and conclusions if band-limited measures are assumed to reflect rhythmic activity without evaluating potential confounding changes in aperiodic activity (Donoghue, Dominguez, et al., 2020). This is important as clinical research has often sought to examine band-specific changes in putative oscillatory activity, some of which may be driven instead by changes in aperiodic activity (Newson & Thiagarajan, 2019). To address this, explicitly separating and measuring both aperiodic and oscillatory measures together is necessary to properly adjudicate which features vary with clinical measures of interest (Donoghue, Haller, et al., 2020). Practically, the recent development of numerous methodological approaches that can separate and measure aperiodic and oscillatory activity has allowed for explicitly examining which features relate to cognitive and clinical correlates of interest (Donoghue & Watrous, 2023).

Collectively, these recent developments have led to a rapid adoption of measures of aperiodic activity in clinical applications, including examining if aperiodic activity may underlie previously reported findings. Much, though not all, of this work is also in the context of seeking ‘biomarkers’, meaning biological measurements that can be used to assist in diagnosis, prognosis, or treatment evaluation of clinical disorders (Aronson & Ferner, 2017; Califf, 2018). In addition to the aforementioned methodological considerations, aperiodic activity is also of interest due to its putative physiological interpretations, which offer the potential for investigating underlying mechanisms of clinical disorders. One such interpretation is its potential relationship with excitatory (E) and inhibitory (I) balance, also referred to as the E/I ratio, whereby a steeper aperiodic component is thought to be related to increased inhibitory activity (Gao et al., 2017).

Overall, this recent work has led to a rapidly expanding literature analyzing aperiodic neuro-electrophysiological features in clinical recordings, across a wide range of different diagnoses (Figure 2A). However, there is as yet no clear consensus on the study of aperiodic neural activity in clinical disorders, including a lack of clarity on if and to what extent aperiodic neural activity relates to clinical disorders; what the key considerations and methodological best-practices are for investigating aperiodic activity in clinical contexts; and to what extent the goals of providing potential biomarkers and/or putative physiological interpretations are being met. To investigate these topics, this systematic review examines this emerging literature, collecting clinically related investigations of aperiodic activity in order to evaluate and integrate information from within and across disorders. In doing so, this review aims to provide brief overviews of key findings from within each disorder, as well as a summary of the current practices, consistencies, and differences across disorders in order to present an overview of the key topics and issues prevalent in this work.

**Figure 2.**
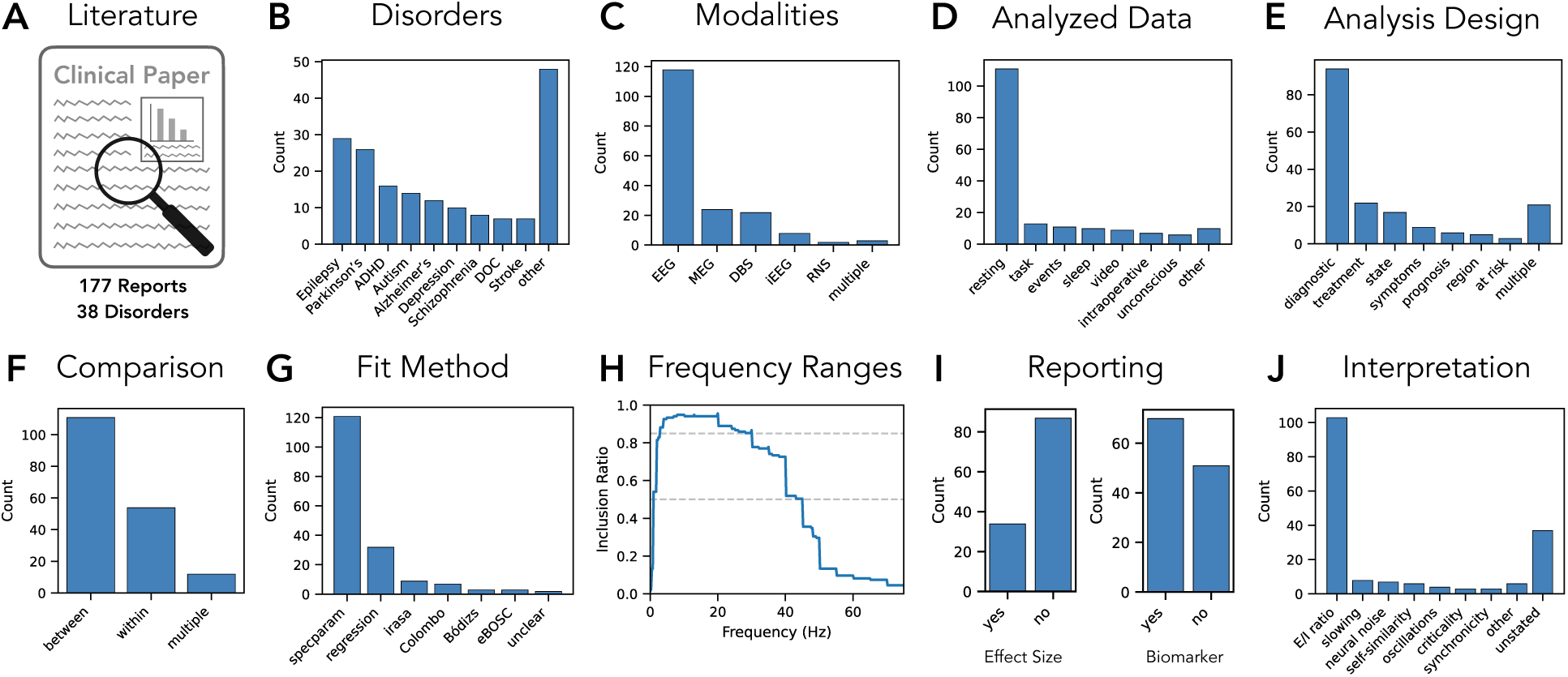
Summary results of the collected literature data. **A)** Literature reporting on the analysis of aperiodic activity in human clinical populations was collected, with 177 reports across 38 disorders found. **B)** Disorders included in the literature dataset. **C)** Recording modalities. **D)** State of the recorded data. **E)** Main analysis design of the study. **F)** Whether comparisons done within or between subjects. **G)** Analysis method used to analyze the aperiodic activity. **H)** Frequency ranges, showing the proportion of reported frequency ranges that include each frequency, for the 135 reports that report a single frequency range. Gray dashed line show thresholds for indicating frequencies included in 50% of analyzed frequency ranges (1-45 Hz) and for frequencies included in 85% of all analyzed frequency ranges (3-30 Hz). **I)** Reported results related information, including whether effect size measure are included, and whether aperiodic activity is discussed as a potential biomarker. **J)** The main stated interpretations of aperiodic activity.

In examining this literature, this review finds that there are many reports of differences in aperiodic neural activity across clinical diagnoses – such that this ubiquity of differences itself raises questions about the specificity and interpretations of such changes. Across this literature, summaries within and across disorders show variability in the results, methods, discussions, and interpretations of aperiodic neural activity in clinical research, and highlight many shared themes across the collective clinical literature. As well as providing mini-reviews of the most studied disorders, by examining the literature across disorders, this approach is able to examine notable patterns across the clinical literature as a whole that may not be identifiable in the much smaller literature of any individual disorder, and identify commonalities in the study of aperiodic neural across clinical research. The main findings, key questions, and shared difficulties of this work are discussed and summarized to make recommendations and provide suggested guidelines for future research investigating aperiodic neural activity in clinical contexts.

## 2. Methods

This project is a systematic review of clinically related work that measures aperiodic activity in neuro-electrophysiological recordings. This literature review included no novel data collection or analysis, nor interaction with any human subjects, and is therefore exempt from ethics review. To introduce key concepts, example simulations were made with the *neurodsp* Python module (Cole et al., 2019) for time-domain simulations and the spectral parameterization (*specparam*; formerly *fooof*) Python module (Donoghue, Haller, et al., 2020) for frequency domain simulations. This project followed the Preferred Reporting Items for Systematic reviews and Meta-Analyses (PRISMA) 2020 guidelines for systematic reviews (Page et al., 2021). The literature collection was done using curated search terms with explicit inclusion and exclusion terms, through a combination of manual search and extraction with the automated ‘literate scanner’ (*lisc*) Python tool (Donoghue, 2019). Automated searches collected references from the Pubmed database, using search terms for each disorder combined with terms related to aperiodic activity. Features of interest were systematically extracted from all included reports, and analyzed within and across disorders, with key themes and discussions topics also collected from across the literature dataset. PRISMA checklists and project materials including lists of search terms, details on the protocols used to extract literature data, information on the collected literature data, and code for simulations, literature collections, and analyses are available in the project repository (https://github.com/TomDonoghue/AperiodicClinical).

Literature searches were done in a two-step process, the first to identify studies examining aperiodic activity in clinical disorders using general terms which was then used to curate a list of disorders that was used in a second phase searching per disorder. Studies that met criteria from either search were included in the analysis. For both phases the following terms were used as search terms for aperiodic neural activity: ‘aperiodic exponent’, ‘aperiodic slope’, ‘spectral exponent’, ‘spectral slope’, ‘1/f slope’, ‘1/f exponent’. In the first phase, these search terms were combined with the ‘or’ search operator and separately searched with the each of the following terms: ‘clinical’, ‘disorder’, ‘disease’, ‘biomarker’, ‘diagnosis’, ‘diagnostic’, ‘treatment’.

From this original search, for any report added to dataset, the name of the disorder was added to a list of disorders that have been examined in relation to aperiodic activity. A second set of searches was then run combining the same aperiodic search terms with each of the disorders, to search for additional reports. The set of disorder terms is included in the Appendix. Further reports were found and added through reference searches of already included reports.

For the literature analyses, reports were included if they examined electrical field recordings (M/EEG, iEEG) from human participants that included patients with a clinical diagnosis as a topic of study and/or at-risk participants with later evaluations for clinical diagnoses. Recordings from electrodes that are part of stimulation devices such as deep brain stimulation (DBS) and implanted random noise stimulation (RNS) devices are also included and referred to by the type of device. Reports were included if they reported an analysis of aperiodic activity as measured from the frequency domain, comparing between clinical group(s) and/or a control group (between subject analyses), and/or if they included analyses within clinical patients, including analyses across clinical events, anatomical areas, or treatment regimens (within subject analyses). Excluded from this review are reports that investigate topics without an explicit diagnosis (e.g. acute intoxication or anesthesia), investigations that only employ time domain methods that cannot be easily compared to frequency domain measures, investigations in animal models, and conference abstracts or conference papers. All reports were screened for inclusion by the author. All reports that met inclusions criteria that were available as published articles or as preprints by December 31^st^, 2024 were included.

To be included, a report had to analyze aperiodic parameters, for example, examining aperiodic activity in relation to a clinical diagnosis, symptomology, and/or treatment. Reports that measured aperiodic activity in the process of examining another feature – for example, being used to normalize measures of neural oscillations without also reporting aperiodic features – were not included. Relevant analyses were restricted to the aperiodic exponent as it is the most analyzed parameter of aperiodic neural activity. No reports were excluded based on only reporting another aperiodic parameter, but some reports do additionally discuss other aperiodic parameters, the details of which are not included here. This review consistently uses the term ‘aperiodic exponent’ (reflecting the χ parameter in the 1/f^x^ formulation), though note that included reports were not required to use this same terminology. For example, the ‘spectral slope’ (b), when referring the slope of the log-log power spectrum, is an equivalent measure (whereby χ = -b), and investigations of this measure are included. For clarity and consistency, in this report all measured values are discussed as exponents (using the conversion above if needed), such that all values are reported as positive, whereby a value of 0 reflects white noise (uniform power across all frequencies), and a value of 1 reflects pink noise (decreasing power across increasing frequencies). With this terminology, an increase in the magnitude of the exponent reflects a steepening of the aperiodic component and a decrease in the exponent reflects a flattening of the aperiodic component.

For each included report, extracted information included clinical information (clinical disorder(s) under study); bibliographic information (title, authors, journal, month and year of publication, digital object identifier (DOI)); dataset information (within or between subject analysis, analysis design, number of patients, number of control participants, ages); recording information (recording modality, what type of data was analyzed (e.g. rest, task, etc.), amount of data (time) analyzed); analysis information (analysis method used to analyze aperiodic activity, the frequency range that was fit, and whether settings and/or goodness of fit measures were reported for the fit method); and results information (the reported results, if and which effect size measures were reported, reported interpretation, and whether this study discusses aperiodic measures as a potential biomarker). In addition, any additional notes about the study were logged, including notes specific to the report and/or relating to discussion topics raised by the report. The complete dataset is available in the project repository, including a full description of how information is coded for each feature.

After collecting and extracting information of interest from the literature, this review sought to synthesize results within individual diagnoses, where possible, as well as collate themes across the entire literature. For clinical diagnoses for which there were at least 5 individual reports (9 disorders), a mini review of the findings for the disorder was performed. These brief overviews had the goal of summarizing the main results and noting the consistency of findings as well as any key discussion topics from within the literature. Note that these within-disorder summaries seek to synthesize results across studies, but are not meta-analyses, and do not include any methods to assess bias or quality, for example weighting of reports by their sample size, or any other quantitative evaluation of the evidence across reports. For diagnoses with fewer than 5 reports each (29 disorders), a synthesis of results within each disorder was not attempted, with this work briefly summarized collectively.

Across the entire literature dataset, patterns were also examined across time, by organizing reports by publication year. As there is an uneven number of reports across different time periods, with all but the recent years having too few reports to compute summary metrics across individual years, publications prior to 2021 were grouped together, and compared to subsequent reports grouped by individual year. From across all reports, key themes were also identified (such as patterns of findings, difficulties of analyses, related discussion points), to examine the commonalities across disorders. Note that while this narrative overview includes some basic quantifications of the literature data (for example, the number of reports with each research design or finding), it overall reflects a largely qualitative overview of the available literature. Finally, based on a combination of the systematically extracted variables as well as the themes identified across reports, a set of recommendations and best practice guidelines are suggested for future work investigating aperiodic neural activity in clinical contexts.

## 3. Results

### 3.1 Results Across All Reports

From the literature search, 177 reports were found that included the examination of 38 different clinical disorders (Figure 2A-B). This literature was published across 94 distinct journals and also included 19 preprints. In total, 10565 clinical patients were reported in this literature (7313 control participants), with a median patient group size of 32 [range: 1-1038] (control: median 36, range: [6-732]). In terms of recording modality, the majority (>60%) of the investigations use EEG (118/177; 67%), with MEG (24; 14%), DBS (22; 12%), iEEG (8; 5%), and RNS (2: 1%) comprising the remainder (Figure 2C). Most investigations analyzed resting state data (111; 63%; Figure 2D). The most common analysis design was investigating diagnosis related differences (95; 54%), with additional designs including treatment response and predicting specific disease states (Figure 2E). Accordingly, most reports employed or included a between-subject comparison approach (111/177 reports; 63%), with the remaining reports also / instead examining within-subject designs (66/177; 37%).

In terms of estimation methods (Figure 2G), the most common method is spectral parameterization (*specparam*; formerly ‘*fooof*’; 121/177; 68%; Donoghue, Haller, et al., 2020), with some usage of other specific algorithms and procedures, including the *irasa* algorithm (9; 5%; Wen & Liu, 2016), the Colombo procedure (7; 4%; Colombo et al., 2019), the *eBOSC* algorithm (3; 2%; Kosciessa et al., 2020), and the Bódizs procedure (3; 2%; Bódizs et al., 2021), as well as a significant number that use a simple linear regression approach (32; 18%). A notable source of variation across reports is the frequency range that is examined – with 63 different specific ranges used within reports that use a single frequency range and an additional 10 reports using multiple different ranges across analyses. However, clustering the related frequency ranges (grouping, for example, ranges 1-40 Hz and 2-40 Hz as similar), shows more commonality across reports. To examine this, the proportion of reported frequency ranges that included each frequency was computed across the full range of frequencies included across all reports (Figure 2H). This shows a relative consistency in analyzing a broad range of predominantly low frequencies, with the frequencies from 1-45 Hz being included in 50% of all analyzed ranges, and the frequencies from 3-30 Hz being included in 85% of all reported ranges. This summary analysis is consistent with 1-40 Hz being the most reported range (13 reports; 7%). Other than this common range, subsets of reports examine shorter ranges (e.g. 1-20 Hz or similar), broader ranges (e.g. 2-55 Hz or similar) and/or ranges starting at higher frequencies (e.g. 20-45 Hz or similar). Notably, 32 (18%) reports have an unclear (not explicitly reported) frequency range.

Additional information was extracted on the reporting of analysis methods. This included noting if the report described method settings and goodness of fit evaluations, which was specifically evaluated for the use of the *specparam* method for which doing so is recommended as best practice (Donoghue, Haller, et al., 2020; Ostlund et al., 2022). This analysis revealed that of reports using *specparam*, only 70/121 (58%) included a partial or full note of settings that were used, and only 34/121 (28%) reported goodness of fit evaluations.

Across the reported results, in diagnostic analyses across groups, 35/110 (32%) reported an increase in the aperiodic exponent in the clinical group, 35 (32%) reported a decrease, 34 (31%) reported no difference, and 6 (5%) did not clearly report the direction of difference. Across all reports (Figure 2I), only 48/177 (27%) included a measure of standardized effect size – most commonly Cohen’s d. In addition, of 110 reports that included an analysis of group differences between clinical and control groups, in only 28 (25%) were the measured exponent values clearly reported. A majority of reports (98/177; 55%) discuss the analyzed features as possible biomarkers (indicating the report discussed aperiodic activity as a potential biomarker, though not necessarily including the conclusion that it is a good biomarker candidate). In terms of the stated interpretations of aperiodic activity (Figure 2J), the most common stated interpretation is E/I ratio (103/177; 58%), with a notable minority not explicitly stating a specific interpretation (37/177; 21%).

The full literature dataset was also examined across time, to examine potential trends and changes. Overall, there is a rapid rise in research (Figure 3A), with the majority of the research conducted in the last several years. To examine features across time while addressing the uneven number of reports, reports from prior to 2021 were grouped together and compared to reports from more recent years. This shows that the number of distinct disorders examined per year has risen (Figure 3B), consistent with the expansion of this literature. The average sample sizes per report is also higher in recent reports (Figure 3C). Methodologically, recent method developments such as *specparam* appear to be replacing the use of simpler linear regression methods (Figure 3D). Examining motivations and interpretations, the discussion of aperiodic activity as a biomarker and/or as a potential marker of E/I ratio appears to be increasing slightly across time (Figure 3E).

**Figure 3.**
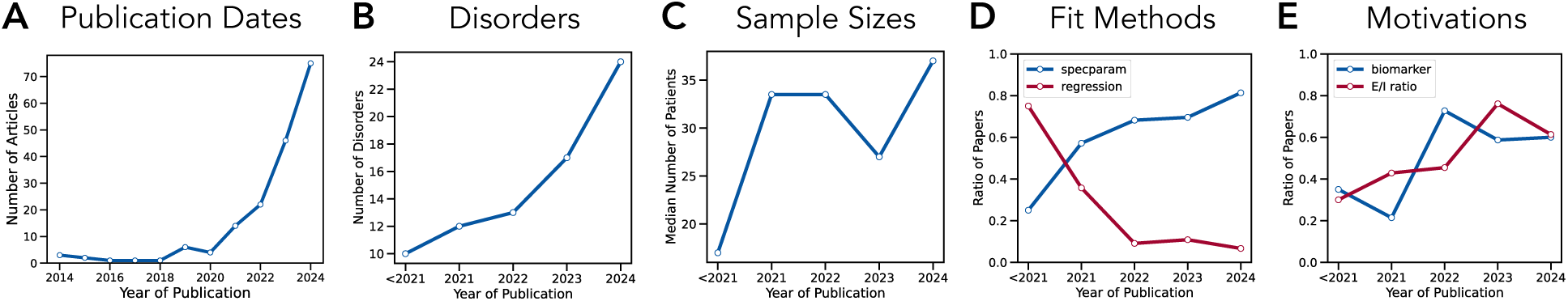
Results Across time. **A)** Publication years of the literature dataset. **B-E)** Properties of the dataset across time, showing **B)** number of disorders studied, **C)** median sample sizes, **D)** fit methods, comparing spectral parameterization (*specparam*) and linear regression methods and **E)** reported motivations and interpretations, reporting proportions of reports interpreting aperiodic activity as related to E/I ratio and discussing aperiodic activity as a possible biomarker. Note that in B-E, time intervals are not equal lengths, as papers prior to 2021 are grouped together (due to the low number of papers per year during this time).

### 3.2 Summaries Within Disorders

In the following, brief summaries of disorders for which there is a sufficient number of reports (>= 5) are presented (9 disorders; total of 129 reports), ordered by the number of reports per disorder. A summary of these disorders including the main findings and key discussions points is reported in Table 1. The remaining 48 reports, covering an additional 29 disorders, are then briefly discussed. The entire set of included reports, including reference information and listings of properties, analyses, and results per report is presented in Table 2.

**Table 1:** Summary of literature in the most studied disorders. For disorders with more than 5 individual reports, a summary across reports was performed. Abbreviations: #: number of reports for each disorder; #/D: number of reports per research design. Modalities are listed in order of occurrence (most used first). Symbols: ⬆ an increase in aperiodic exponent was reported; ⬇ a decrease in aperiodic exponent was reported; **∅** no difference in aperiodic exponent was reported; Δ changes or differences (across listed topic) were reported and discussed.

| Disorder | # | Modalities | Design | #/D | Main Findings | Discussion Topics |
| --- | --- | --- | --- | --- | --- | --- |
| Epilepsy | 29 | EEG, iEEG, MEG, DBS, RNS | state treatment region | 17<br>5<br>2 | ↑ prior/during seizure (13↑; 2↓; 2 both)<br>↓ with treatment (4↓; 1∅)<br>↑ in seizure onset zones | Δ across events / brain states<br>Δ across anatomical locations<br>Δ across frequency ranges |
| Parkinson's | 26 | DBS, EEG, MEG | diagnostic treatment symptoms | 10<br>7<br>2 | ↑ clinical vs. control (8↑; 1↓; 1∅)<br>↑ with medication (4↑; 4∅)<br>inconsistent ~ w symptom (2 yes; 3 no) | Δ across modalities / groups<br>Δ across symptom measures |
| ADHD | 16 | EEG | diagnostic treatment | 15<br>4 | ↓ clinical vs. control (9↓; 3↑; 3∅)<br>varies with treatment - inconsistent | Δ across age / development<br>Δ with treatment / condition |
| Autism | 14 | EEG, MEG | diagnostic symptoms | 9<br>3 | inconsistent (5∅; 1↑; 4↓)<br>relates to symptoms, idiosyncratically | Δ across age / development<br>Δ with specific symptoms |
| Alzheimer's | 12 | EEG, MEG | diagnostic region | 9<br>3 | inconsistent (5∅; 2↑; 2↓)<br>report region specific differences | Δ across etiology / progression<br>Δ across anatomical locations |
| Depression | 10 | EEG, DBS | diagnostic treatment | 5<br>4 | ↓ clinical vs. control (3↓; 2∅)<br>↑ with treatment (3↑; 1↓) | Δ across modalities / groups<br>Δ across anatomical locations |
| Schizophrenia | 8 | EEG, MEG | diagnostic | 8 | inconsistent (4∅; 2↑; 1↓; 1 both) | Δ across states (tasks)<br>Δ across anatomical locations |
| DOC | 7 | EEG | diagnostic prognosis | 4<br>3 | ↑ clinical vs. control (3↑; 1↓)<br>↓ ~improved clinical scores | Δ across etiology / progression<br>Δ in analysis methods / ranges |
| Stroke | 7 | EEG, MEG | diagnostic region | 6<br>4 | ↑ clinical vs. control (5↑; 1∅)<br>↑ over affected hemisphere (4↑) | Δ across age / progression<br>Δ across anatomical locations |

**Table 2:**
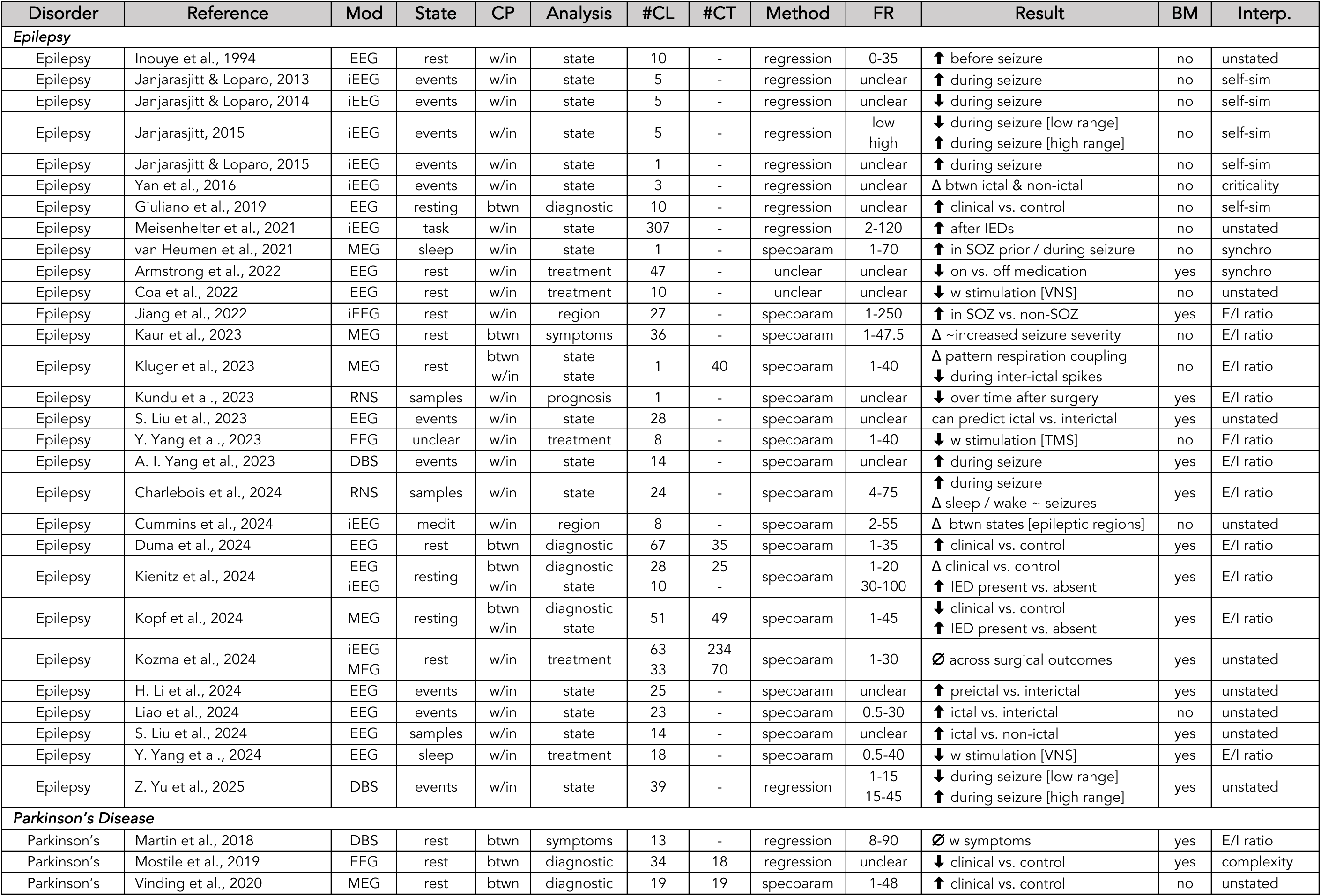
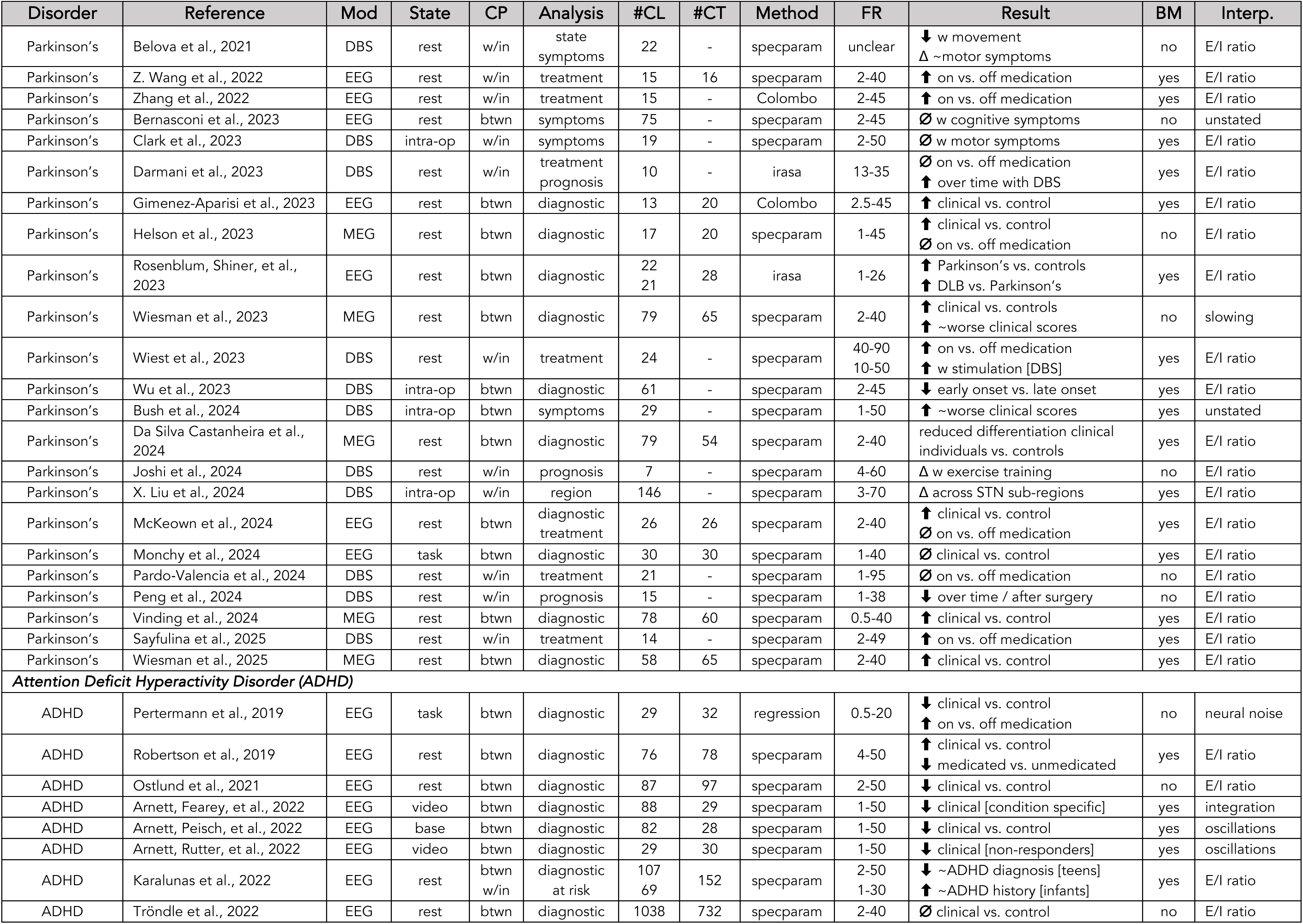
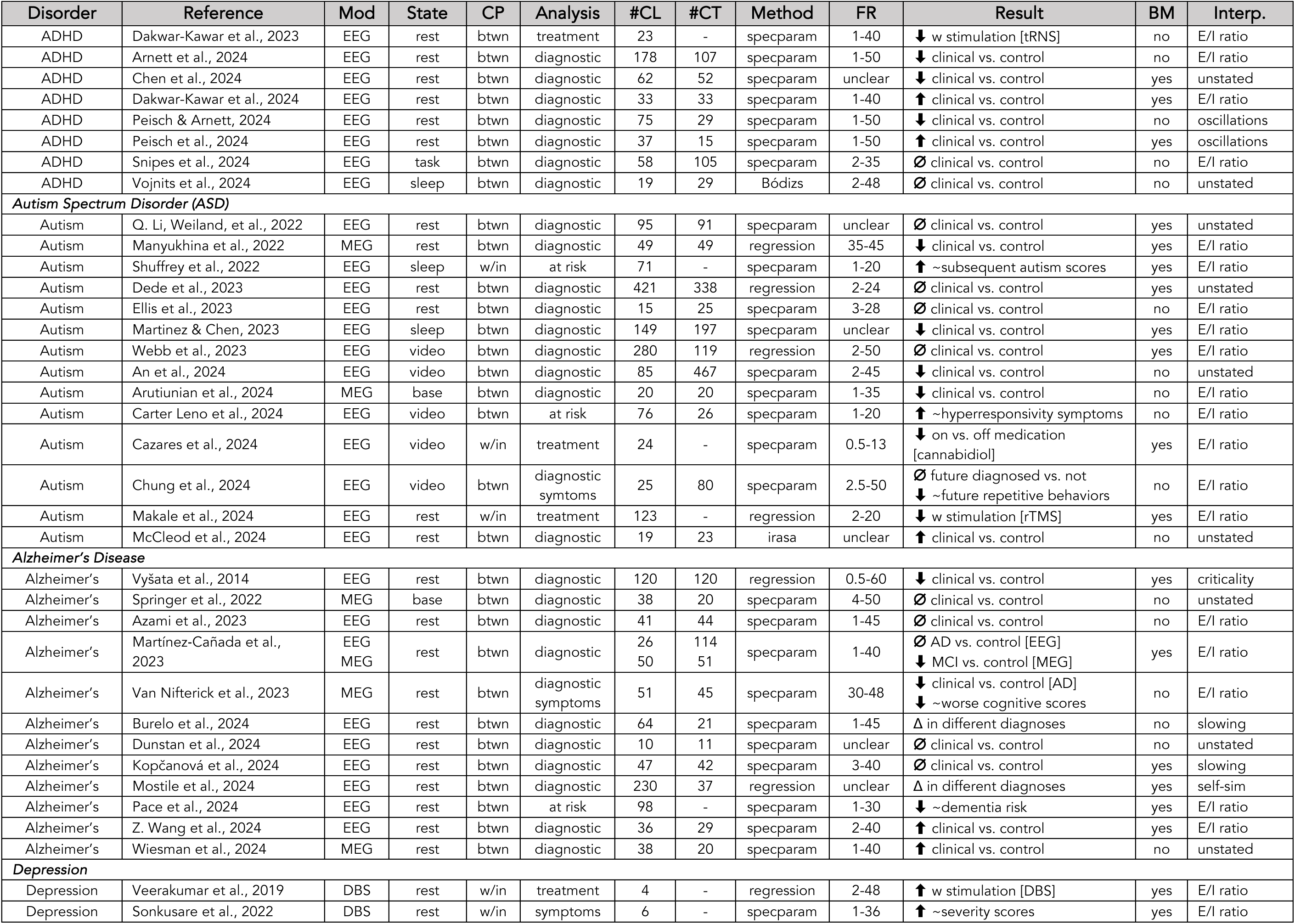
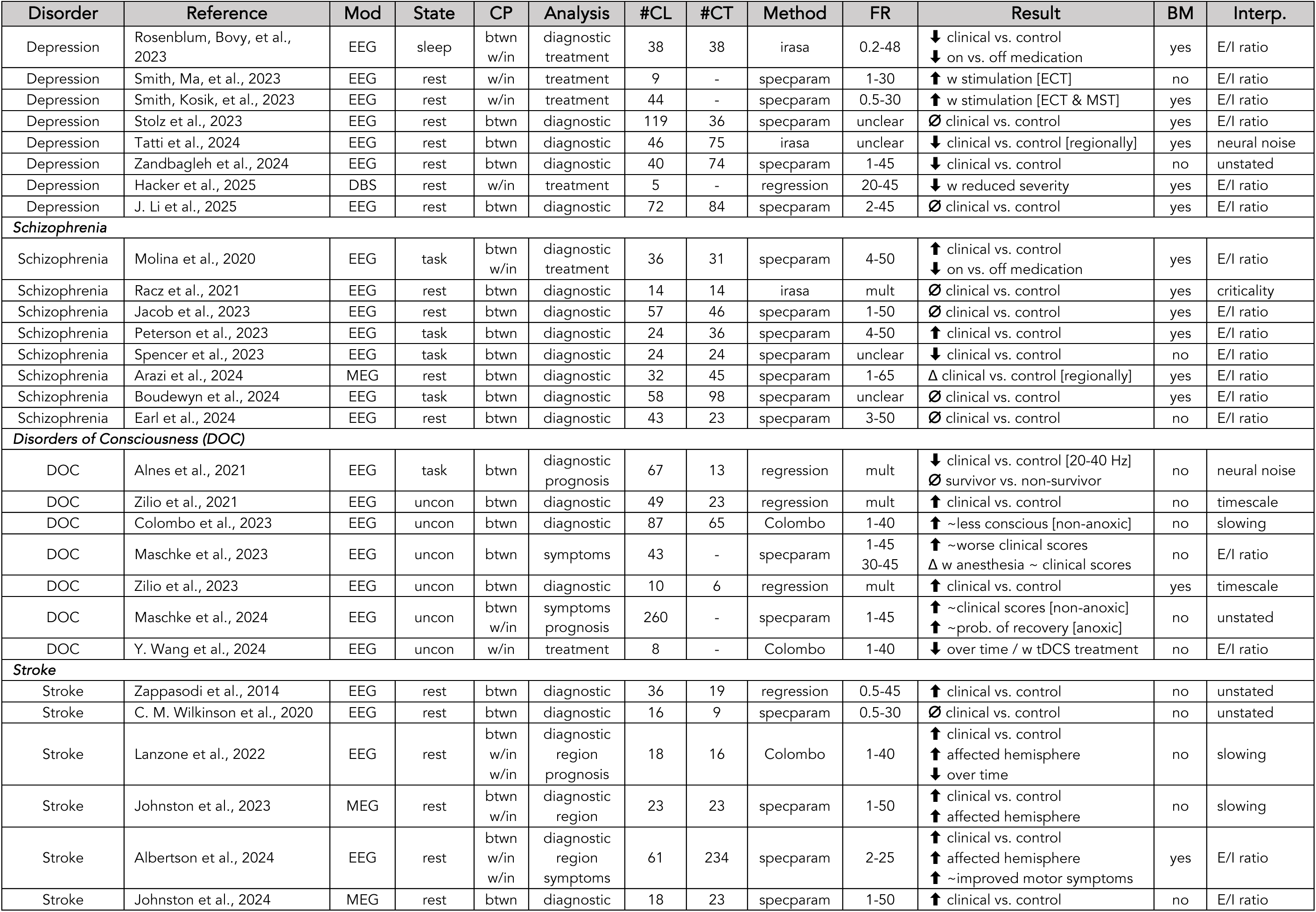
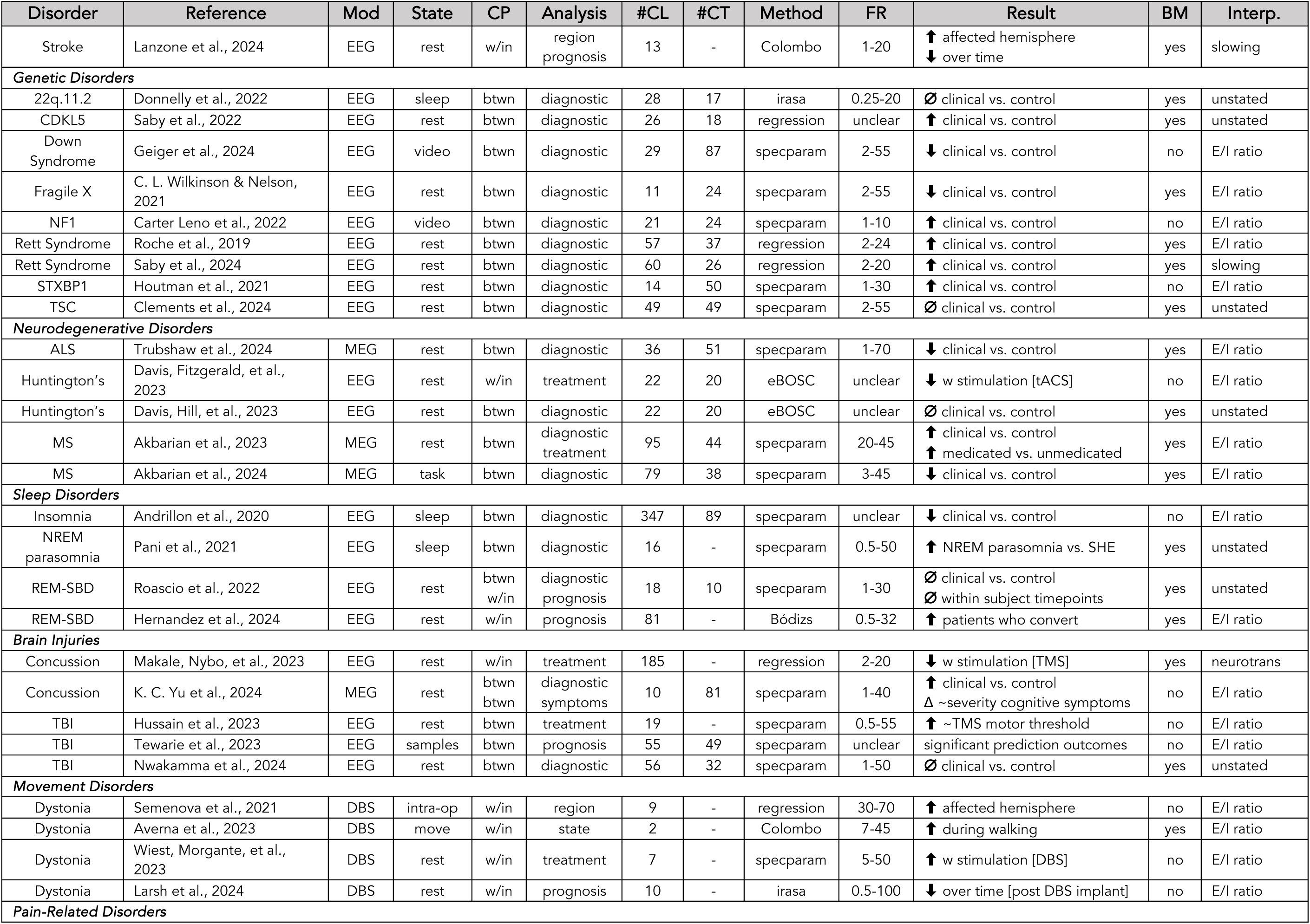
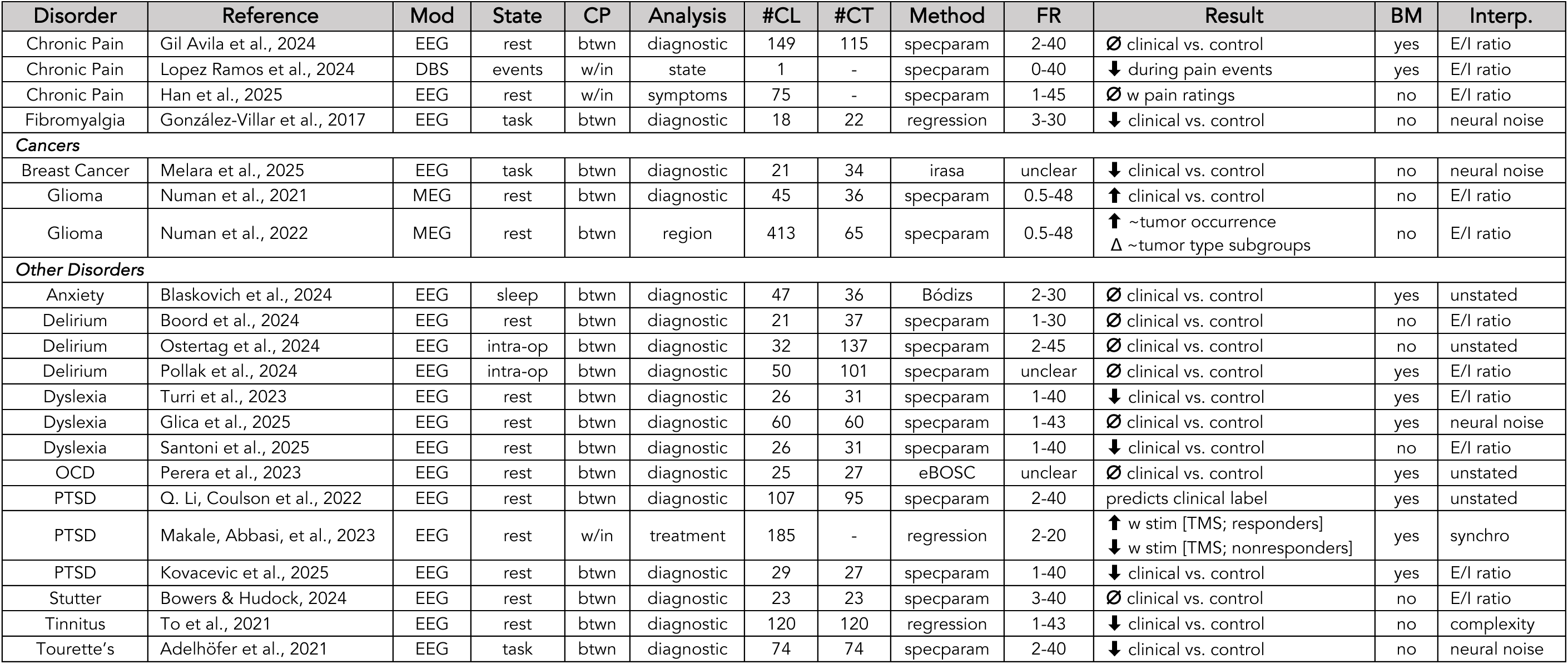
Dataset of all reports investigating aperiodic neural activity with clinical populations. All reports identified and included in the literature dataset, organized by disorder. Each report has the following fields: *Disorder*: the clinical diagnosis under investigation in each report. ***Reference***: the bibliographic reference for the report. Note that publication year listed for the reference may be different from date used to evaluate eligibility (e.g. a report may be accepted and available in a different calendar year than the reference year once included in an issue). ***Mod*** (Modality): the recording modality of the data. References to stimulating devices (e.g. DBS, RNS) refer to recordings from electrodes that are part of stimulating devices. ***State***: the recording state of the data. ***CP*** (Comparison): the analysis design as within (w/in) or between (btwn) subjects. ***Analysis***: the main analysis design of the report. ***#CL***: the number of clinical participants. ***#CT***: the number of control participants (if relevant). ***Method***: the analysis method used to measure aperiodic activity. ***FR*** (Fit Range): the frequency fit range, in Hz, the method was applied to. ***Result***: the main aperiodic exponent related result(s) of the report. Arrows refer to a finding of an increased (⬆; steepening) or decrease (⬇; flattening) of the aperiodic exponent; ∅ refers to a finding of no difference in the aperiodic exponent or no relationship to the exponent; Δ refers to a change or difference in the measure; and ∼ refers to an association (e.g. correlation) between the exponent and the reported measure. ***BM*** (Biomarker): whether the report discusses aperiodic activity as a potential biomarker. ***Interp*** (Interpretation): the main interpretation of aperiodic activity discussed by the report. **Abbreviations** - *Disorder column*: 22q.11.2: 22q.11.2 Deletion Syndrome; ADHD: attention deficit hyperactivity disorder; CDKL5: CDKL5 Deficiency Disorder; MS: multiple sclerosis; NF1: Neurofibromatosis type 1; OCD: obsessive-compulsive disorder; PTSD: post-traumatic stress disorder; REM-SBD: REM Sleep Behavior Disorder; TBI: traumatic brain injury; TSC: tuberous sclerosis complex. *State column*: base: baseline; intra-op: intraoperative; medit: meditation; move: movement; mult: multiple; uncon: unconscious. *Modality column*: DBS: deep brain stimulation; EEG: electroencephalography; iEEG: intracranial EEG; MEG: magnetoencephalography; RNS: responsive neurostimulation. *Result column*: AD: Alzhiemer’s dementia; DLB: Dementia with Lewy Bodies; ECT: electro-convulsive therapy; IED: interictal epileptiform discharges; MCI: mild cognitive impairment; MST: magnetic seizure therapy; rTMS: repetitive TMS; SHE: sleep-related hypermotor epilepsy; SOZ: seizure onset zone; STN: subthalamic nucleus; tACS: transcranial alternating current stimulation; tDCS: transcranial direct current stimulation; TMS: transcranial magnetic stimulation; tRNS: transcranial random noise stimulation; VNS: vagus nerve stimulation. *Interpretation column*: neurotrans: neurostransmission; self-sim: self-similarity; synchro: synchronicity

#### 3.2.1 Epilepsy

Epilepsy is the most examined clinical disorder (29 reports; median # patients: 14 [range: 1-307]), as well as the one with the most longstanding interest, including most of the oldest reports in the dataset. In contrast to most other disorders, investigations are mostly not oriented around between-subjects comparisons examining diagnostic differences, but instead largely relate to within-subject analyses of different states (seizure detection, e.g. comparing differences between ictal (during) and interictal (between) seizure events). Within this work, there is a high degree of consistency across reports, with pre-ictal and ictal activity tending towards having a larger aperiodic exponent – though note that there are idiosyncrasies relating to which event categories and timepoints are examined. Similarly, regional comparisons tend to find a greater aperiodic exponent in seizure onset zones, as compared to control regions. A small number of reports also suggest a decrease in the aperiodic exponent with treatment (across different kinds of treatment). Interestingly, in epilepsy there is some work explicitly comparing different frequency ranges, including reports of different findings across different frequency ranges. Collectively, the work in epilepsy is broadly consistent with temporally and regionally specific changes in aperiodic activity systematically relating to seizure activity.

#### 3.2.2 Parkinson’s

Parkinson’s disease is the second most studied disorder in the collected literature, with 26 reports (median # patients: 23 [range: 7-146]), across M/EEG and DBS, investigating diagnostic, prognostic, treatment, or symptom-related hypotheses. For diagnostic comparisons, there is a fair amount of consistency – of 10 clinical to control group comparisons, 8 report an increased exponent in Parkinson’s, 1 reports a decreased exponent, and 1 reports no difference. Investigations comparing between on and off medication include 4 reports that find an increased exponent when on medication, and 4 reports finding no difference, suggesting a tendency for medication to increase the exponent. Symptom comparisons include 3 reports finding no relationship to cognitive or motor symptoms, and 2 reporting a correlation between increased exponent and worse clinical scores. The variability in the later comparisons may relate to differences across recording modalities, treatment details, and symptom measures, with some reports also investigating and discussing regional differences. A common theme across this literature is that the motivation for measuring aperiodic activity was often noted as including the goal of better isolating beta oscillations, which are also implicated in Parkinson’s disease. Reports that investigated both aperiodic and periodic features generally report that they both relate to disease status, and that separating the components assists with investigating the relationships of each to clinical features. Collectively, this literature suggests a generally consistent relationship of an increased (steepened) exponent relating to Parkinson’s disease with somewhat less consistency when examining to what extent this relates to treatment status and symptom measures.

#### 3.2.3 ADHD

The study of attention-deficit/hyperactivity disorder (ADHD) is focused on reports using EEG to compare clinical populations to control groups to examine diagnosis-related differences in aperiodic activity (16 reports; median # patients: 68 [range: 19-1038]). The diagnostic results across these reports are somewhat variable, with 9 reports finding a lower aperiodic exponent in the ADHD group, 3 reports finding a higher exponent, and 3 finding no difference. What is emerging across this research is the variable nature of aperiodic activity in this population – results are reported to interact with demographics such as age, treatment status, and task condition of the recording. Age appears to explain some of the differences across reports, as well as patient group characteristics, including several reports demonstrating an effect of medication status on aperiodic exponent, which is not limited to acute status and can persist after drug washout. There is thus far only minimal work that evaluates aperiodic activity in relation to symptoms. Collectively, the literature on ADHD suggests a complex pattern of differences in aperiodic activity and suggests that the heterogeneity of the populations under study – including variation in age, treatment status, and symptomology – contributes to variability that needs to be considered and addressed in order to examine differences in more targeted sub-groups.

#### 3.2.4 Autism

The investigation of autism (14 reports; median # patients: 74 [range: 15-421]) includes mostly EEG investigations comparing clinical patients to control patients, including some work on at-risk populations comparing measured parameters to future diagnoses. The results across these reports are variable – while 4 report a decreased exponent in autistic individuals or those who are later diagnosed as autistic, 1 reports an increase, and 5 report no difference. Two at risk studies report an increased exponent relating to later clinical measures, but using different measures. This variability perhaps relates to the heterogeneity of autism – 2 reports examining relationships to symptom scores reported relationships of aperiodic activity to specific symptom measures, even in the absence of group level diagnostic differences. There is also a high variability of age ranges across reports, including distinct developmental stages ranging from infancy to adulthood, and age-related effects / confounds are discussed in this literature. Overall, the available evidence in autism therefore suggests that there is not a clear and consistent pattern of aperiodic activity across all diagnosed individuals, though there may be more nuanced relationships between subgroups of patients and/or to specific symptoms of the disorder.

#### 3.2.5 Alzheimer’s Disease

The study of Alzheimer’s Disease includes 12 reports (median # patients: 49 [range: 10-230]) that examine differences between clinical and non-clinical groups in MEG and EEG analyses, all of resting or baseline data. The results of comparisons between patients with Alzheimer’s and control groups are quite variable, including 5 reports finding no differences, 2 reporting a lower exponent in the clinical group, and 2 reporting a higher exponent in the clinical group. One key consideration that may relate to the variable findings are differences in disease etiology and progression, as analyses comparing different disease states (e.g. mild cognitive impairment vs. Alzheimer’s) and/or including comparisons to other dementia-related diagnoses report differences between distinct clinical groups. This suggests differences in aperiodic activity may be dynamic across disease progression and/or specific to disease etiology. Several investigations also reported region-specific differences, such that differences in modality and analyzed areas may contribute to the reported differences. Collectively, the study of Alzheimer’s dementia does not suggest a clear and consistent difference in such patients (as compared to control), though the broader comparison of dementia suggests potential differences that may be specific to progression, etiology, and/or anatomical regions.

#### 3.2.6 Depression

The work on depression (10 reports; median # patients: 39 [range: 4-119]) includes a mixture of work examining diagnostic comparisons, treatment related responses, and symptom-related investigations. Notable across this literature is the variation in recording modality, with a mix of EEG and DBS, which have significant differences in the anatomical locations and sources of the recorded data. Many of the examined treatments are stimulation based, which are generally consistent in reporting increases in the aperiodic exponent with stimulation treatment. This is also broadly consistent with 3 diagnostic studies that reported a decreased exponent in patients as compared to controls – though another 2 studies reported no difference between groups. However, 2 reports examining clinical severity scores instead reported that a flattening of the exponent correlated with decreased clinical severity. Overall, the literature in depression includes a high degree of variation of modalities and designs that are difficult to compare (not only due to modality differences themselves but the likelihood that there may be differences between patients who are eligible for invasive treatments such as DBS as compared to those on standard care), with a suggestion overall that depression may in some cases show a decreased aperiodic exponent and that treatment for depression may increase the aperiodic exponent.

#### 3.2.7 Schizophrenia

The study of aperiodic activity in schizophrenia (8 reports; median # patients: 34 [range: 14-58]) is mostly with EEG, and is by majority focused on examining diagnostic differences. Reported diagnostic results are overall inconsistent, with 2 reporting an increased exponent in schizophrenic patients as compared to controls, 1 reporting a decreased exponent, and 4 reporting no difference. In addition, 1 study investigating differences across anatomical locations found a pattern of differences including both increases and decreases across the cortex, suggesting that there may be anatomical variation in the diagnostic effect on the exponent.

Despite the consistency in recording modality and subject demographics (all young adults), there are considerable differences in the analyzed data, with multiple different tasks being analyzed, potentially also relating to the differences in results across reports. There is thus far limited evidence on the effect of pharmacological treatment, with one study reporting a treatment-related decrease of exponent in schizophrenic patients, and limited investigation of symptoms or cognitive measures. Overall, the literature in schizophrenia does not suggest a clear and consistent difference across all patients, with potential impacts of anatomy, the recording state of the data, treatment status, and symptom dimensions currently unclear.

#### 3.2.8 Disorders of Consciousness

In disorders of consciousness (DOC) research (7 reports; median # patients: 49 [range: 8-260]), key research questions include examining whether aperiodic activity can help dissociate between different diagnoses (e.g. vegetative state vs. locked in syndrome) and/or predict future recovery. Reports examining the aperiodic exponent are quite consistent in suggesting an increased exponent is related to disorders of consciousness, and that a lower exponent is related to better clinical scores and treatment response. Notably, however, recent investigations have emphasized that this is not a ubiquitous finding across all DOC patients, with notable differences across different etiologies, in particular comparing between anoxic and non-anoxic patients. Across this work, there have been multiple different frequency ranges, analysis methods, and patient groups examined, such that the precise details of which specific measures vary in which specific groups is still a topic of ongoing research. Collectively, this work suggests that when addressing differences in etiology of DOCs, the aperiodic exponent has a fairly consistent relationship to both diagnosis and clinical measures.

#### 3.2.9 Stroke

The study of stroke includes 7 reports (median # patients: 18 [range: 13-61], which mostly investigate diagnosis related differences, as well as notable investigations of regional differences, and if and how measures of the aperiodic exponent relate to prognosis. In comparing between patients and control, there is a general consistency in finding a higher exponent in patients, as found in 5 reports, with 1 additional report finding no difference. This is also consistent with analyses from 4 investigations examining regional differences which all report an increased aperiodic exponent over the hemisphere of the stroke. In addition, 2 reports with longitudinal recordings find a decrease of the aperiodic exponent over time. It is as yet unclear from the current investigations if and how measures of the exponent relate to clinical scores and prognosis. Overall, the work in stroke is consistent in finding diagnostic related differences, with variation across anatomy (as it relates to the stroke), and across time since the clinical event.

#### 3.2.10 Other Disorders

Beyond the reports summarized thus far, an additional 48 reports across a further 29 disorders were collected in the literature dataset, including diagnoses relating to sleep disorders, genetic disorders, neurodegenerative diseases, brain injury related disorders, movement disorders, pain related disorders, cancers, and other psychiatric and non-psychiatric disorders (median # patients: 29 [range: 1-413]). The number of distinct diagnoses – including 17 diagnoses with a single report each – further emphasizes the breadth of aperiodic-related investigations in clinical contexts. Across this additional literature, the majority of investigations (23/29) report at least one significant difference between groups and/or a treatment related effect of aperiodic activity. Largely due to the large number of examined disorders (and the small number of reports per disorder) there are no clear patterns to note – with diagnostic differences being reported as both increases and decreases, as well as multiple and variable relationships reported across treatment-related, prognosis-related, and regional comparisons. Several reports also include multiple different clinical groups that are compared together. Common discussion points include the heterogeneity of clinical groups and variation across diagnoses, treatments, modalities, and regions, which is overall consistent with discussion points raised within individual disorder evaluations. Combining across all disorders, 32 out of the 38 diagnoses included in this investigation include at least one report claiming a relationship between aperiodic activity and disease status, treatment, or symptoms – showing that differences in aperiodic activity are a common finding in clinical disorders.

## 4. Discussion

This literature review examined investigations of aperiodic neural activity in clinical contexts, summarizing 177 reports across 38 distinct clinical diagnoses. The consistency of results across disorders varies, with the most studied disorders of epilepsy and Parkinson’s having arguably the most consistency in their results, as well as a high degree of consistency in neurological disorders such as stroke and DOC. By comparison, psychiatric disorders appear to generally have less consistent results, consistent with the longstanding difficulty in identifying consistent biomarkers in psychiatric conditions (García-Gutiérrez et al., 2020; Venkatasubramanian & Keshavan, 2016). Most of the included disorders included too few individual reports to examine the results across reports. By examining across the different disorders, several themes emerged across multiple different disorders – of heterogeneity, notable covariates, limitations in current methodological practice, and discussion of interpretations of aperiodic activity – that can be used to develop recommendations for best practices to pursue further work evaluating aperiodic neural activity in clinical contexts.

### 4.1 Sources of Variation in Measures of Aperiodic Activity in Clinical Work

A key pattern is the heterogeneity within and across disorders – related but different diagnoses, subject demographics, disease etiologies, symptom clusters, or brain states during recording have been shown to have differing findings. This was noted in reports of autism, ADHD, DOC, dementia, depression, and schizophrenia – disorders in which there are variable findings across reports. This heterogeneity has different forms – for example, in the case of DOC and dementia, findings suggest different diagnoses, etiologies and/or disease progression can have different relationships with aperiodic activity; in ADHD and potentially autism there appears to be differences across age / developmental stages; in schizophrenia as well as ADHD there is some suggestion of differences in task conditions relating to differences in results; and differences in findings in depression may relate to difference in the subject populations that participate in different treatments. Ultimately, across many different diagnoses, the pattern that is emerging is that it appears to be common for clinically-related differences in aperiodic activity to be moderated by subject demographics, clinical etiology, disease progression and/or symptom clusters – motivating these considerations as key factors for designing robust analysis strategies – rather than differences in aperiodic activity reflecting a simple binary difference of with vs. without diagnoses.

Another key theme is the effect of treatment, whereby direct investigations of treatment-related effects as well as investigations comparing clinical to control groups (and not aimed at examining treatment response) have often noted an effect of treatment (pharmacological or otherwise) on measured aperiodic activity. The effect of pharmacological medication is most persistently discussed in reports of ADHD and Parkinson’s disease, as well as some work in schizophrenia and depression, and includes evidence that such differences can extend beyond acute drug effects such that they are not necessarily addressed by research designs that use a drug washout period prior to recording. There are also numerous investigations showing differences in non-pharmacological treatments, including invasive and non-invasive brain stimulation. These findings emphasize the importance of employing approaches that can seek to delineate differences due to the disorder vs. differences due to treatment.

Across investigations and disorders, there is also the topic of anatomical specificity. In disorders such as epilepsy, Parkinson’s, and stroke, for which there are often hypotheses about focal origins of disordered activity, there is evidence for regional specificity in aperiodic differences (for example, within vs. outside the seizure onset zone in epilepsy, across different regions of the basal ganglia in Parkinson’s, and between hemispheres in stroke patients). In some cases, this may also relate to differences in aperiodic activity between cortical and sub-cortical locations (Bush et al., 2024). Notably, many of the surveyed investigations average results across electrodes / regions, including some reports which average all electrodes across the whole head. Further research is needed to evaluate if and when such averaging is appropriate, and/or if it may be sub-optimal due to potentially masking region-specific differences, and/or increased susceptibility to artifact sources such as muscle noise in peripheral electrodes. An additional consideration is potential differences based on different modalities – while this review included reports from across multiple different recording modalities, if and how potential differences such as their spatial specificity and differences in sensitivity relate to measurements of aperiodic features is currently unknown. Future non-clinical work on the spatial properties of aperiodic activity would be of great benefit – including examining the spatial properties of aperiodic activity, modality related differences, and best-practices for if and how to average results across channels and regions. Within clinical applications, future work may benefit from more systematically considering anatomical variation within and between groups.

As well as the relative consistency in epilepsy and Parkinson’s, stroke and disorders of consciousness (when controlling for differences in disease etiology) also have quite consistent results. This suggests increased consistency of findings in diagnoses for which there is relatively greater understanding of hypothesized regions of interest and physiological underpinnings and/or may relate to increased detectability of differences in neurological conditions in which there is injury to or degeneration of brain tissue. The relative preponderance of invasive recording modalities in epilepsy and Parkinson’s may also relate to increased consistency, likely due to the regional specificity and increased signal-to-noise ratio of invasive recordings, but also, speculatively, perhaps due to the increased similarity of patient populations who meet criteria as surgical candidates. By comparison, in most psychiatric disorders (for which, broadly speaking, there are not robust physiological descriptions), and where extracranial recordings are more common, the results thus far do not generally support clear diagnosis related differences but rather more complex interactions of differences in aperiodic activity that may vary with other patients characteristics (e.g. age, treatment status, etiology, disease progression, etc.). Collectively, across neurological and psychiatric conditions, general themes suggest multiple dimensions of variability, including clinical heterogeneity, impacts of treatment effects, and differences across anatomical regions.

### 4.2 Methodological Related Discussion Points

One of the key motivating factors for studying aperiodic activity, as stated explicitly in many of the examined reports, is for methodological validation of whether reported differences between clinical and control groups reflect oscillatory or aperiodic features. Recent reviews and methodological investigations have noted that many reported clinical findings that have often been interpreted as relating to rhythmic coordination of neural activity could instead reflect aperiodic activity, for example a common pattern of increased power at low frequencies and decreased power at high frequencies (Newson & Thiagarajan, 2019) and/or a change in measured ‘band ratios’ of power across low and high frequency ranges (Donoghue, Dominguez, et al., 2020; Finley et al., 2022). As established across the examined literature, there is now evidence that in many disorders there is indeed evidence for differences in aperiodic activity. In some cases, these findings have been evaluated to potentially ‘explain away’ previous reports on predefined oscillation bands, for example in ADHD where differences in aperiodic activity may explain previous reports of differences in theta / beta ratio. It’s important to emphasize, however, that there is not a general answer to whether oscillatory and/or aperiodic features relate to clinical diagnoses, and it needs to be evaluated on a per case basis. Results in Parkinson’s and Alzheimer’s, for example, have established that measuring and controlling for aperiodic activity assists in isolating oscillatory activity and can improve measured associations between aperiodic-adjusted oscillatory features and clinical features of interest, even in reports that also find associations with aperiodic activity. This also emphasizes the importance and relevance of evaluating and reporting null results for aperiodic activity, as this may be instrumental for investigating other features, such as neural oscillations (Donoghue et al., 2022). Collectively, the findings here are consistent with noting the importance of separating and measuring aperiodic and oscillatory features together, to best adjudicate which features relate to measures of interest.

The recent emphasis on investigating aperiodic features and separating them from oscillatory activity is reflected in multiple recently developed analysis methods (Donoghue & Watrous, 2023), many of which were used in the examined literature. Broadly speaking, while comparisons of methods such as *specparam* and *irasa* have found that these methods are similar in their performance, simple linear regression approaches, which are quite common in the examined literature, are generally worse performing (Donoghue et al., 2024) and should be avoided in future work. This review also focused on reports that explicitly measure and interpret the aperiodic exponent as measured from the power spectrum. Another aspect of aperiodic activity is the aperiodic offset, reflecting shifts of the entire spectrum, which is thus far less studied, but can also offer physiological insight (Manning et al., 2009), and should be further investigated in future work to evaluate if and how this feature may relate to clinical diagnoses in addition to and/or differently from the aperiodic exponent. There are also numerous other measures of complexity, entropy, and similar measures that highly overlap with spectral measures of the aperiodic component (Donoghue et al., 2024). Collectively this suggests that beyond this rapidly growing research on frequency domain measures of aperiodic activity, many other reports using related measures likely reflect similar and/or overlapping dynamics in the data, and future work should seek to compare to and integrate these findings.

Regardless of which analysis method is used, there needs to be consistent and clear protocols and reporting guidelines to ensure consistent quality control and reporting such that results can be further integrated and meta-analyzed. In the collected literature, one of the most variable aspects of the analysis is the examined frequency range. This is an important source of variation, as some of the inconsistencies in reported results may reflect differences in the measured frequency range of the data. Notably, if electrophysiological recordings were strictly 1/f distributed, it would not matter what range was analyzed, as all ranges would be self-similar. However, in practice such data is not strictly 1/f (hence the term 1/f-like), and the presence of oscillatory peaks, artifacts sources, filters, and various other features can lead to the examined range having significant impact on the measured parameters. The variability in frequency range also relates to considering the interpretations of aperiodic activity – for example, the relationship to E/I ratio is proposed to be most strongly related to a specific range of frequencies (Chini et al., 2022; Gao et al., 2017). In addition, aperiodic components can have ‘knees’, where there is a change in the 1/f scaling, which appear as a bend in the log-log spectrum (Gao et al., 2020). There is some evidence that variations in knees, if not accounted for, may underlie measured changes in the exponent when analyzing short frequency ranges (Ameen et al., 2024). Future work should investigate and develop best practices for applying well-motivated and consistent frequency ranges, as well as considering how this relates to potentially different forms of aperiodic activity and putative interpretations.

Measuring aperiodic neural activity also raises its own set of methodological questions. Most of the reports involved resting state recordings with relatively short amounts of data (median: 5 minutes, range: [20 seconds - 40 minutes]). The evidence thus far (in MEG) suggests that aperiodic estimates are stable with about one minute of data (Wiesman et al., 2022), suggesting current practice in terms of amount of data is likely adequate, though more work, including with clinical populations, is needed on this topic. Another consideration for the potential use of aperiodic neural activity as a potential biomarker is the test-retest reliability of such measures. There has recently been a series of investigations examining test-retest reliability of aperiodic parameters in healthy adult subjects (McKeown et al., 2024; Pathania et al., 2021; Pauls et al., 2024; Tröndle et al., 2023), which all reported intra-class correlations above 0.7, and some much higher, reflecting high reliability. Notably, the aforementioned investigations were done in healthy, adult participants. In a clinical context, investigations of children with autism have reported good, though lower, intra-class correlations in the range of 0.5-0.7 (Levin et al., 2020, Webb et al, 2023). Future work should continue to validate test-retest reliability scores for aperiodic exponent estimation across broader age ranges (including children), and across more clinical populations.

Methodologically, a key goal for continued work on aperiodic neural activity in clinical contexts should be the development of normative measures of key features across large populations of clinical and non-clinical participants that can be used to compare to clinical groups. There is already some research on this topic, including evaluations of aperiodic parameters across large, primarily non-clinical datasets (>1000 participants) that help to establish norms (Hernandez et al., 2024; Tröndle et al., 2023). It’s also important to consider that many clinical populations are young (infant and early childhood, for developmental disorders) or older adults (for late-in-life diseases). Such age groups may not be well-represented in non-clinical work that often examines healthy young adults, thus requiring dedicated work to examine such populations, with some existing large-sample investigations of young (McSweeney et al., 2023) and old (Cesnaite et al., 2023) populations. In addition, there has recently been an investigation of aperiodic activity in a large dataset of clinical recordings that sought to establish clinical norms (Leroy et al., 2025). Collectively, this work is starting to provide information on expected values and ranges that future clinical work can compare to, with further work needed for establishing normative values for clinical and non-clinical populations.

### 4.3 Interpretations of Aperiodic Neural Activity in Clinical Reports

Being able to record population activity, especially non-invasively, and infer circuit properties is a key goal – but also a difficult problem – for cognitive, computational, and clinical neuroscience (Ahmad et al., 2022; Cohen, 2017; Martínez-Cañada et al., 2021; Pesaran et al., 2018). One such circuit property of interest, E/I balance, is hypothesized to relate to numerous clinical disorders (Ferguson & Gao, 2018; Foss-Feig et al., 2017; Gao & Penzes, 2015; Selten et al., 2018; Sohal & Rubenstein, 2019). The interpretation of aperiodic neural activity as a potential marker of E/I balance is clearly a driving factor of clinical work, being by far the most common stated interpretation in the reviewed clinical reports – though note that different potential interpretations of aperiodic activity are not necessarily mutually exclusive, so this does not imply other interpretations are invalid or irrelevant.

Evidence for the link between the aperiodic exponent and E/I balance comes from computational models (Chini et al., 2022; Gao et al., 2017; Lombardi et al., 2017; Trakoshis et al., 2020) and empirical demonstrations of patterns of aperiodic activity across sleep, anesthesia, and task engagement that are broadly consistent with the expected pattern given this interpretation (Colombo et al., 2019; Gao et al., 2017; Lendner et al., 2020; Waschke et al., 2021). However, there is still a relative lack of direct evidence from physiological manipulations that clearly establishes this link, and the available evidence is not definitive. Invasive animal model recordings including optogenetic stimulation show that increasing or reducing inhibitory activity leads to changes in activity consistent with changes in the aperiodic exponent (Chini et al., 2022), as do pharmacological approaches that increase inhibition (Gonzalez-Burgos et al., 2023). However, subsequent work with pharmacological and optogenetic manipulations found that while an increase in inhibition does lead to an increase in the exponent, an increase in excitation does not reliably decrease the exponent (Salvatore et al., 2024). Manipulations of dopamine have also been reported to flatten the aperiodic exponent in animal models (Kim et al., 2022; Valencia et al., 2012), though such effects may not be as robust in human patients (see Parkinson’s section on medication effects). A flattening of the exponent in response to dopamine manipulation could be interpreted as an increase in excitation – however, that dopamine, a neuromodulator, influences the exponent in such a way when more direct manipulations of excitatory neurotransmitters do not is overall not entirely consistent with the E/I balance interpretation of the aperiodic exponent.

Collectively, the evidence motivates that while physiological manipulations do impact aperiodic activity – consistent with its biological relevance – they do not do so entirely consistently with the predictions of simple models of excitation and inhibition. While driving inhibition more consistently evokes the expected effect on aperiodic activity, the effect of excitation is less clear, with a lack of demonstration of expected effects of increased excitation through direct manipulations of excitatory activity and/or transmitters, while flattening is observed with neuromodulators such as dopamine. Notably, E/I balance, while clearly a powerful and important concept, is also a complex one, with further specification needed in specific cases to define whether differences in E/I balance are expected in terms of the number of neurons, the amount of different neurotransmitters, and/or the activity patterns across neurons (Ahmad et al., 2022). This can also be seen in the reviewed clinical literature whereby discussions of changes in E/I balance include cell death (e.g. in DOC & stroke), neurotransmitter availability (e.g. ADHD & Parkinson’s), and differences in connectivity (e.g. autism & epilepsy).

In addition, there are other proposals for underlying factors that can relate to the measured aperiodic exponent, which may or may not be consistent with changes in E/I. Some of the reviewed reports discuss ‘neural noise’ or similar ideas such as ‘synchronicity’ which posit that changes in the aperiodic component relate to changes in the correlation structure of underlying neural activity (Voytek et al., 2015). Relatedly, theoretical descriptions of 1/f activity such as in relation to criticality, as mentioned by some of the reviewed reports, describe how complex systems such as the brain evolve in their dynamics through time and what functional properties this may have – notions that have also been considered in relation to clinical disorders (Zimmern, 2020). It has also been shown that oscillation damping, reflecting changes in the amplitude dynamics of oscillatory components, can affect the aperiodic exponent (Evertz et al., 2022). While these different approaches and frameworks are not necessarily at odds with descriptions of E/I, they also offer other perspectives for considering such changes in neuro-electrophysiological recordings.

Combining the above discussion of the empirical data, with this broader discussion of E/I, emphasizes some key points about the prominent E/I balance interpretation of the aperiodic exponent, including that i) differences in E/I balance (and therefore aperiodic exponent, in so far as it reflects E/I) can reflect multiple, distinct underlying physiological changes; ii) not all changes in E/I balance have the expected impact on measured aperiodic exponent, suggesting it is not a clear one-to-one mapping; and iii) aperiodic activity is a coarse, global measure, and observed relationships between it and E/I do not preclude that other, non-E/I related changes may also impact the measured exponent. Collectively, this suggests that given the current evidence, a change in aperiodic activity may or may not reflect a change in E/I balance, the lack of a change in the aperiodic exponent does not necessarily imply the lack of a change in E/I balance, and the same change in aperiodic exponent can likely arise from different underlying changes in E/I related or non-E/I related features. As such, a change in aperiodic activity, by itself, should not be strongly interpreted as a direct marker of E/I balance.

While this lack of a clear general relationship between the aperiodic exponent and E/I complicates simple interpretations of changes in aperiodic activity, it also helps explain how seemingly similar patterns of differences can be seen across such a range of disparate disorders.

The causes of the noted changes in aperiodic activity across a broad range of clinical disorders are likely underdetermined – varying across disorders – and may relate to variable underlying differences in E/I balance, from various sources, and/or to other aspects of neural function. Drawing from the clinical literature to better contextualize these different potential underlying changes across disorders may lead to better understanding the putative sources of aperiodic activity. Notably, recent work is seeking to address this complexity, for example, in modeling approaches that can examine more detailed biophysical interpretations of changes in power spectrum structure (Bloniasz et al., 2025). Overall, the current research suggests that more nuanced interpretations of concepts such as E/I balance, motivated by more detailed modelling of changes in the power spectrum, are needed – with future work needed to integrate recent advances into the discussion and interpretation of changes in aperiodic activity seen in clinical disorders.

### 4.4 Aperiodic Activity as a Potential Biomarker

Given these methodological and scientific considerations, it’s worth revisiting the idea of the aperiodic exponent as a potential biomarker, as discussed by many of the included reports. The term ‘biomarker’, while rapidly increasing in use in the literature, is used in variable ways and is often poorly defined (Aronson & Ferner, 2017; Califf, 2018). Notably, few of the included reports explicitly define what is meant by ‘biomarker’ in their respective usages. In psychiatry in particular, there have been longstanding attempts, but little progress in developing objective biomarkers, which has been the topic of much debate (García-Gutiérrez et al., 2020; Venkatasubramanian & Keshavan, 2016). The exploration of aperiodic activity as a potential biomarker should learn from this background – presenting clear and specific definitions that clarify how the term is being used, how it is envisioned as contributing to clinical practice, and doing so considering the history of similar attempts and detailing how known problems and limitations will be addressed.

In most cases in the collected literature, the term biomarker is used in relation to examining diagnostic related differences between groups. It’s important to note that examining mean group difference between groups – the most common approach taken – is insufficient to evaluate a measure as a diagnostic biomarker, as this approach does not consider the variability within each group (Loth et al., 2021). Relatedly, the frequent lack of standardized effect size measures limits the consideration of findings as potential biomarkers. Additionally, almost all reports examine only one diagnosis, but across the literature many disorders are reported to have differences in aperiodic activity – with 32/38 disorders included in this investigation including at least one report of a relationship between aperiodic activity and disease status, treatment, or symptoms. This lack of specificity of differences in aperiodic activity, as well as diagnostic comorbidity and the difficulty this poses for prediction from electrophysiological features (Langer et al., 2022), suggests that while non-normative measures of aperiodic may reflect a general indicator of abnormal activity, it is not currently established that aperiodic activity has the reliability or specificity to serve as a biomarker to assist with differential diagnoses.

Importantly, many of the mentions of biomarkers were not diagnostic related, but rather relate to symptom scores and/or within-subject prediction of future state, for example, in discussions of treatment response (to pharmacology or stimulation) and/or in examining prognosis. Such cases may be more promising for the potential use of aperiodic measures as markers for examining treatment response and/or tracking disease progression – with more longitudinal research needed for such cases. Additionally, addressing the sources of heterogeneity, as mentioned across the surveyed literature, will also be key for discussions of biomarkers. This could include, for example, examining aperiodic activity in particular regions and/or in relation to particular symptoms in appropriately curated subgroups of patients with similar disease progressions and/or etiologies which could potentially reveal currently elusive levels of reliability and specificity.

Given that much of the reviewed literature motivates the study of aperiodic activity due to its potential as a measure of E/I balance and/or as a putative biomarker, both of which have limitations, it is worth considering the future of examining aperiodic activity in clinical work. First, as discussed, measuring and controlling for aperiodic activity is important for adjudicating which features vary in neuro-electrophysiological recordings, and so measures of aperiodic activity should still be included as part of methodological best practice. In doing so, while our current understanding of the physiological underpinnings of aperiodic neural activity is incomplete, the available evidence does support that it relates to properties of underlying circuits — even if our current simple models do not fully capture how it does so — such that ongoing and future work on the physiological underpinnings of aperiodic activity holds promise for contributing to further understanding the neurophysiology of neurological and psychiatric pathologies (Bloniasz et al., 2025). Additionally, the limitations highlighted here should not eclipse that many of the included analyses do report promising findings regarding diagnostic, treatment, and/or state related differences in aperiodic activity that may well be clinically useful, and work within and across different disorders is productively highlighting considerations to improve the robustness and interpretability of future studies such that it’s potential use as biomarker for certain applications is still plausible. Notably, most of the literature included in this review reflects work across just a few years as methods and ideas have rapidly developed, setting the stage for future work to build on this foundation, address current limitations, and further evaluate the utility of measuring aperiodic neural activity in clinical disorders.

### 4.5 Recommendations for Future Work

To best be able to engage in productive future work on aperiodic activity in clinical disorders, this systematic review suggests some key recommendations based the clinical findings and the ensuing discussion of the methodological and scientific themes. These recommendations are also summarized in a checklist (Table 3), drawing from the summarized literature to assist with developing best practices for continued work in this area. At the level of individual reports, it is important to establish clearly defined goals for the investigation – whether it is, for example, an explicit search for a biomarker for diagnosis and/or treatment (including clarifications of what is meant by biomarker) and/or a study to examine potential mechanisms (including clarification on how the exponent is being interpreted). In designing experiments, collecting data samples, and creating analyses, it’s important to consider topics that have arisen in this literature – for example, confounds of age / development, variation across etiology / severity, medication-related changes, regional differences, etc. – to ensure that research designs are best suited to examine questions of interest.

**Table 3:**
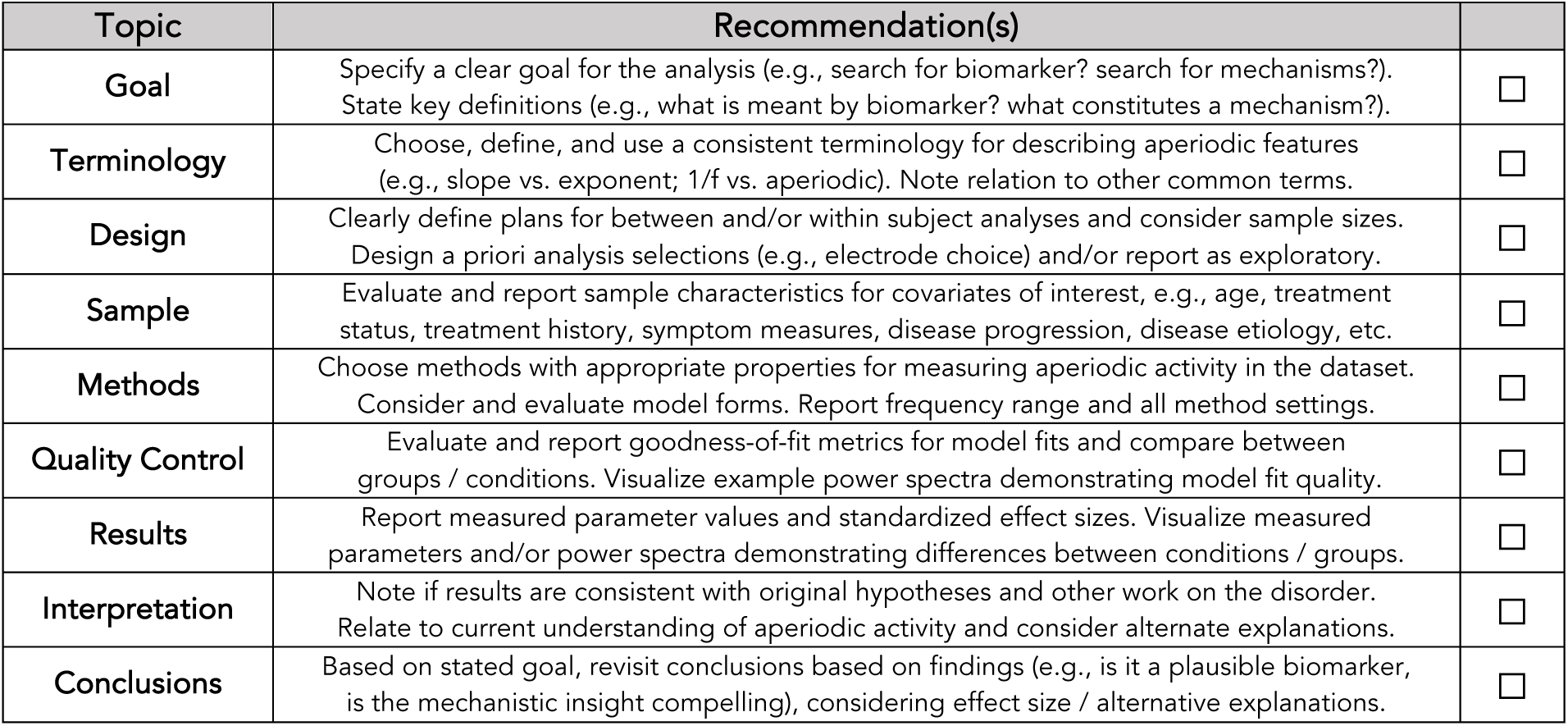
Checklist of recommendations for clinical investigations of aperiodic activity. Each topic includes a summary of recommendations to check / consider for research investigating aperiodic activity in clinically related investigations. Note that not all recommendations are relevant for all research designs. See main text for further details.

One aspect that would benefit from improved consistency and clarity is terminology. This review included reports that discuss aperiodic activity described in multiple ways, including the terms ‘aperiodic exponent’ (χ) and ‘spectral slope’ (b). Either description is valid and they are functionally equivalent (whereby χ = -b). There are however reports that use both, sometimes interchangeably, which can be confusing and create interpretational issues – since slope values are negative and exponent values positive, the report of a decrease / increase of the aperiodic parameter is ambiguous if the unit / value is not clearly established (since a decrease of 2 to 1 exponent values is a flattening, whereas a decrease of −1 to −2 of slope values is a steepening, though this is also complicated by some reports seeming to discuss increases / decreases of slope magnitude rather than of actual value). In cases where actual values are not reported and/or results are not visualized, the actual results can be quite unclear. Terms such as “flattening” and “steepening” are useful as they are unambiguous regardless of the measured quantity. Reports are recommended to consistently employ a single, consistent description of aperiodic activity, and clearly and consistently report data values and directions of changes.

Methodologically, the reviewed literature includes multiple different analysis methods and high variability in the reporting standards for describing the use of these methods. Most notably, the analyzed frequency range must be properly reported, as well as any settings for the chosen method. There is also a lack of reporting of goodness-of-fit measures that can help to establish the quality of the model fits, and limitations in the reporting of measured parameter values. In designing and applying analyses, researchers should consider possible anatomical differences and choose included electrodes and regions of interest accordingly, avoiding approaches such as averaging across all electrodes which may neglect regional differences, and may be more susceptible to artifacts. For quality control of measures, use of model fitting methods such as *specparam* should include the evaluation and reporting of goodness-of-fit measures, including potential group differences in model fit quality and checking for and potentially excluding outliers. It is also useful to examine and provide example individual and/or group average model fits, which can be used to demonstrate model fit quality and to visualize differences that are quantified by the models.

In terms of results reporting, when possible, it is useful to report parameter values per group / condition. This allows for evaluating if the values are in an expected range, as more reports of normative values become available, and makes such values available for inclusion in future meta-analyses and comparisons to future datasets. Where appropriate, the calculation and reporting of standardized effect sizes of group / condition differences can help to evaluate the magnitude of effects and discriminability of patients based on such features, beyond only reporting if there are significant differences. Reports should also include clear statements on how individual parameters relate to diagnostic and/or treatment related features of interest ensuring, for example, that prediction-based analyses that mainly report accuracy include information on the direction of differences between groups / conditions. In reporting the results, interpretations, and conclusions, reports should make a clear connection between the analysis results and the original goals and hypotheses, while considering the magnitude of effects, the current status of research on interpretations of aperiodic activity, and possible alternate explanations.

To best support the investigation of aperiodic activity in clinical conditions, the above noted recommendations should be supplemented by the continued development of standardized guidelines and protocols for investigating aperiodic neural activity. There also needs to be consistent communication between clinical and non-clinical work – with clinical investigations having both much to contribute to the broader understanding of aperiodic neural activity and its interpretations, and much to gain from non-clinical methodological, cognitive, and physiological investigations. In particular, the research field as a whole will benefit from work continuing to pursue large-scale norming studies, with and without clinical subjects, to establish clearer definitions of normative values for aperiodic activity; methodological work continuing to evaluate method-related best-practices, the impact of methodological choices such as frequency range and different model forms, and relationship(s) between distinct methods; physiological work probing the underlying circuit mechanisms that drive changes in aperiodic activity; and ultimately, the development of standardized and evidence-based protocols for best-practices to measure aperiodic activity.

### 4.6 Limitations

There are some limitations to the approach taken in this systematic review. Perhaps most notably is the high-level overview of a large number of reports covering a broad range of investigations in this study. This approach – reducing reports to a single or small number of briefly summarized key results – necessarily ignores many details of the investigations, limits the nuances of the results that are discussed, and may miss notable details that could help explain findings and patterns within and across diagnoses. The inclusion of many different disorders, and many different research designs (e.g. diagnostic, treatment-related, symptom-related, etc.) also precludes more systematic and quantitative meta-analytic approaches, such that the overall approach here includes multiple mini-reviews within disorders as well as a largely qualitative overview across the entire literature. The inclusion of different recording modalities, including both intra- and extra-cranial methods, adds variability to the dataset which may explain some differences across reports and disorders, and precluded a detailed consideration of analysis procedures such as pre-processing steps and the handling of artifacts which vary across modalities, and which may impact measures of aperiodic activity. While extensive literature searches were undertaken for the current review, it is possible that some relevant reports were missed, in part due to the breadth of the potentially relevant literature, and variability in the terminology used across different reports.

In addition, the organization of reports based on a single diagnosis does not fully reflect the details of a small number of reports that included multiple different and/or comorbid diagnoses, nor does it integrate information across different but similar or related diagnoses. Within individual disorders, summaries of findings were not weighted by sample size or quality metrics, nor was it evaluated if studies may have any overlaps in the analyzed data (e.g. drawing from the same open datasets) to evaluate if reports can be considered independently of each other. Accordingly, this investigation should only serve as a general summary across the clinical work at large, whereby future work should be dedicated to more fully examining the details of investigations within individual disorders and/or related groups. This review was also performed by a single investigator and as such did not include consensus methods across multiple reviewers assessing elements such as study selection and inclusion. This review also includes qualitative summaries reflecting the selection, interpretation, and prioritization of discussion points from across the literature, which could potentially be considered differently by other authors and should be further discussed by others in future work.

## 5. Conclusion

Aperiodic neural activity has rapidly emerged as a feature of interest in clinical research, as evidenced by the rapid rise in the number of reports across time. This research has made many contributions, including methodological work evaluating which neural features relate to clinical disorders and providing potential markers to track treatment and prognosis; and scientific work, probing potential physiological interpretations of disease-related changes or differences in neural function. However, some caution is warranted, as across fields and disorders there are common issues and discussion points that often complicate the conclusions; overlapping findings across disorders that suggest a lack of specificity in the results; and ongoing discussions of the interpretations of aperiodic activity that complicate it’s straight-forward interpretation as a biomarker and its potential relationships to underlying circuit activity. Future work can hopefully use this interim check-in on the status of clinically related reports of aperiodic neural activity to guide future investigations on how to examine and interpret this feature in clinical work.

## Appendix: Literature Search Terms

The following details the search terms used for the literature collection. For further details on how these terms were used, see the Methods section. For the use of these search terms in automated literature searches, see the Project Repository.

**Aperiodic activity search terms (used for all searches):**

’aperiodic exponent’, ‘aperiodic slope’, ‘spectral exponent’, ‘spectral slope’, ‘1/f slope’, ‘1/f exponent’.

**Phase 1 search terms for clinically related reports on aperiodic activity:**

’clinical’, ‘disorder’, ‘disease’, ‘biomarker’, ‘diagnosis’, ‘diagnostic’, ‘treatment’.

**Phase 2 search terms for reports per disorder:**

“parkinson’s”; ‘epilepsy’, ‘seizure’; ‘ADHD’, ‘attention deficit hyperactivity disorder’; ‘autism’, ‘ASD’; ‘alzheimers’, ‘dementia’; ‘disorders of consciousness’, ‘coma’, ‘locked-in’; ‘depression’, ‘MDD’, ‘major depressive disorder’; ‘schizophrenia’; ‘stroke’; ‘dystonia’; ‘TBI’, ‘traumatic brain injury’; ‘dyslexia’; ‘glioma’; “huntington’s”; ‘multiple sclerosis’; ‘PTSD’, ‘post traumatic stress disorder’; ‘REM sleep behavior disorder’; ‘rett syndrome’; ‘22q.11.2’; ‘ALS’, ‘amyotrophic lateral sclerosis’, “Lou Gehrig’s disease”; ‘anxiety’; ‘CDKL5 deficiency disorder’; ‘chronic pain’; ‘concussion’; ‘down syndrome’; ‘fibromyalgia’; ‘fragile X’; ‘insomnia’; ‘NF1’; ‘NREM parasomina’; ‘OCD’, ‘obsessive compulsive disorder’; ‘STXBP1’; ‘tinnitus’; ‘tourette’; ‘tuberous sclerosis complex’; ‘delirium’; ‘stutter’; ‘cancer’;

**Exclusion terms, used to ignore unrelated literature:**

’acid’, ‘protein’, ‘ion’, ‘enzyme’, ‘ultrasound’, ‘cancer’, ‘halide’, ‘spectroscopy’, ‘iodide’, ‘tissue’.

## Data Availability

The literature data collected in this systematic review are available in the project repository and linked from the manuscript.

https://github.com/TomDonoghue/AperiodicClinical

## Acknowledgements

The author would like to thank Jonathan K Kleen, Sydney Smith, and Bradley Voytek for helpful comments on this manuscript.

